# Short-term animal product restriction alters metabolic profiles and modulates immune function

**DOI:** 10.1101/2025.05.23.25328246

**Authors:** Eleni M. Loizidou, Aikaterini Palaiokrassa, Samuel Assiedu, Alexandros Simistiras, Petros Barmpounakis, Stavros Glentis, Maria Anezaki, Kristina V. Bergersen, Iosif Chatzimichalis, Eleni Kotsanopoulou, Ioannis Kontoyiannis, Nikolaos Demiris, Pavlos Rouskas, Nikolaos Scarmeas, Mary Yannakoulia, Mihalis Verykokakis, Meera G. Nair, Konstantinos Rouskas, Antigone S. Dimas

## Abstract

Diet shapes immune function and disease susceptibility, yet the underlying mechanisms remain poorly defined. Here, we investigated the immunometabolic effects of short-term dietary restriction of animal products in a unique group of Greek individuals who alternate between omnivory and animal product restriction for religious reasons. We profiled clinical biomarkers and immune parameters during both dietary states, alongside a control group of continuously omnivorous individuals. Dietary restriction was associated with reductions in total and non-high-density lipoprotein cholesterol, urea, creatinine, alanine aminotransferase, and gamma-glutamyltransferase, related to improvements in cardiovascular, renal and hepatic function. We also found a striking 73% reduction of normal range C-reactive protein levels. Immune profiling revealed reductions in non-classical monocytes, CD56⁺ Natural Killer cells, and CD8⁺ memory T cells, accompanied by increased levels of anti-inflammatory cytokine IL-10, suggesting a shift toward a less inflammatory immune state. Our findings demonstrate that short-term restriction of animal products rapidly improves metabolic and immunological health-related markers and may lower risk of chronic inflammatory disease. These insights highlight the translational potential of short-term dietary interventions in altering health-related risks.

## Introduction

Dietary intake influences all physiological processes, including regulation of the immune system. Diet in industrialized countries, frequently referred to as the “Western diet”, is typically rich in red meat and highly processed foods (Veldhoen & Veiga-Fernandes, 2015) and has been shown to contribute to the incidence of chronic inflammatory disorders and risk for other diseases (Afshin et al, 2019). Over the past decades, dietary patterns that partially or wholly exclude animal products have become popular due to their association with lower risk of chronic diseases, including obesity, cardiovascular disease, type 2 diabetes, rheumatoid arthritis, and cancer (Dinu et al, 2017; Key et al, 2022; Kjeldsen-Kragh et al, 1991; Yannakoulia & Scarmeas, 2024; Zhang et al, 2023). To date, few comprehensive studies have addressed effects of plant-based diets on health, but collectively, findings from multiple studies have shown that individuals who follow plant-based diets are likely to have lower body mass index (BMI), lower blood pressure and lower levels of blood lipids, insulin and glucose compared to individuals following conventional omnivorous diets (Dinu et al, 2017; Key et al, 2022). Additionally, plant-based diets have been associated with lower blood levels of urea, creatinine, gamma-glutamyltransferase (γ-GT), and inflammatory markers, such as C-reactive protein (CRP) (Tong et al, 2021). Although most effects of plant-based diets appear to be beneficial for human health, there are also unfavourable consequences including lower bone mineral density, increased risk of bone fractures and of haemorrhagic stroke, compared to diets including meat (Key et al, 2022; Tong et al, 2020).

Effects of plant-based diets on disease outcomes are likely exerted in part, through modulation of the immune system, but few studies in humans have explored the underlying mechanisms. A four-week vegan dietary intervention led to reductions in the number of total leukocytes, neutrophils, monocytes and platelets compared to a diet rich in meat (Lederer et al, 2020), while a two-week vegan diet intervention resulted in increased numbers of activated T helper cells, Natural Killer (NK) cells, and upregulation of pathways associated with antiviral immunity (Link et al, 2024). Furthermore, we have shown that short-term, periodic dietary restriction (DR) of animal products results in substantial immunometabolic reprogramming, including reductions in plasma levels of proteins linked to inflammation (Rouskas et al, 2025). Overall, studies focusing primarily on model organisms, have highlighted that responses linked to T cells and cells of the innate immune system can be shaped by various forms of mild or transient DR (Collins & Belkaid, 2022), but this this remains largely underexplored for plant-based diets in humans (Link et al, 2024).

In the present study, we carried out comprehensive profiling of health-related blood biomarkers and of immune system cells for a unique group of healthy adults from Greece who alternate between omnivory and DR of animal products for religious reasons for 180-200 days annually, in a highly consistent and temporally structured manner. The high consistency of the dietary pattern and the predictability of the shift between omnivory and animal product restriction, renders this study comparable to an interventional study. We compared effects of this dietary pattern to a continuously omnivorous control group profiled in parallel and found that short-term DR was associated with substantial decreases in blood lipids and markers of renal and liver function. DR was also linked to a 73% decrease in normal-range CRP levels and to a profile suggesting reduced cardiovascular disease (CVD) risk. Furthermore, following DR we found decreased frequencies of non-classical monocytes, CD56^+^ NK cells and CD8^+^ memory T cells, along with an enhanced response in the anti-inflammatory cytokine IL-10. We also uncovered seasonal changes in the responses of pro-inflammatory cytokines IL-8, MCP-1 and TNF-α. Our findings suggest beneficial effects on health and improved inflammatory profiles of even short courses of animal product restriction, that are likely to drive better outcomes in chronic inflammatory disease.

## Results

### Population sample and characteristics

Participants who met FastBio (religious <u>Fast</u>ing <u>Bio</u>logy) study criteria (Rouskas et al, 2025) (**Appendix Text S1**) belonged to one of two dietary groups: individuals who voluntarily alternate between omnivory and DR of animal products for religious reasons in a highly predictable and structured manner (periodically restricted, PR group N=200), and continuously omnivorous control individuals (non-restricted group, NR group N=211) **(Fig.1)**. PR individuals, who practice DR of animal products for a total of 180-200 days annually, constitute a unique study group given the high consistency of their dietary pattern and the predictability in switching between omnivory and restriction (**Appendix Text S2**). This voluntary dietary behaviour renders our study similar to a long-term dietary intervention study. Participants were profiled at two time points: T1 in autumn, covering a period of omnivory for both dietary groups, and T2 in early spring, covering three-to-four weeks of DR for PR individuals, during Lent. Typically, during this period, PR individuals undergo restriction of protein (as a proportion of total energy intake), mainly driven by abstinence from almost all sources of animal protein, which is not accompanied by a decrease in total energy intake (Georgakouli et al, 2022; Sarri et al, 2004).

**Figure. 1.**
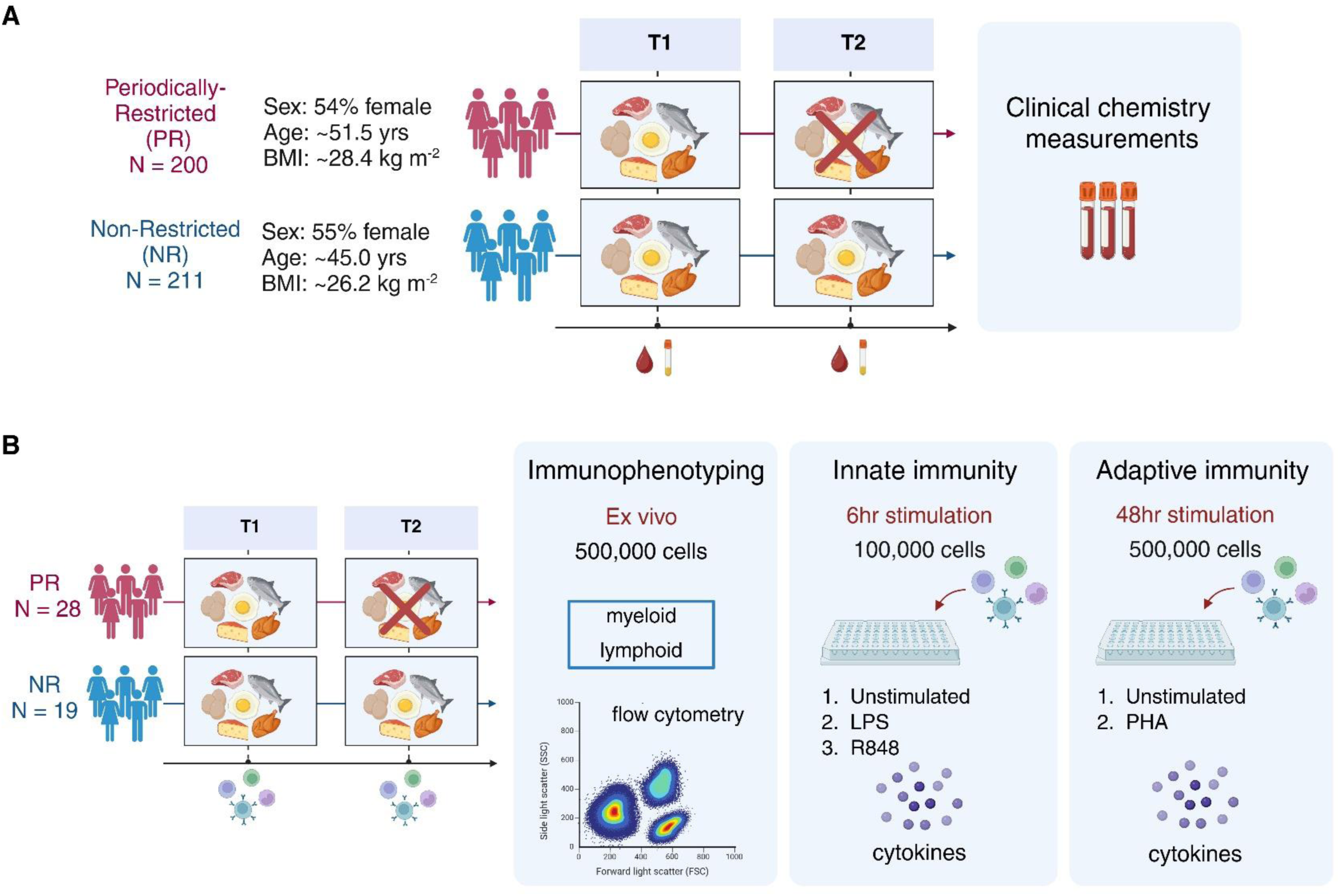
Study design. **A.** Two hundred participants practicing periodic restriction of animal products (PR group, red) and 211 continuously omnivorous, non-restricted participants (NR group, blue) were profiled for blood biomarkers and complete blood counts (CBC) at two time points. T1: both dietary groups were on an omnivorous diet. T2: the PR group had practiced animal product restriction for 3-4 weeks. **B.** For a subset of participants at both time points, we isolated PBMCs and: i) performed ex vivo profiling of immune cells through flow cytometry (left panel), ii) quantified production of cytokines by PBMCs at 6 hours following stimulation with LPS and R848 to capture effects on innate immunity (central panel), and iii) quantified production of cytokines by PBMCs at 48 hours following stimulation with PHA to capture effects on adaptive immunity (right panel).

On average, PR individuals were older (PR: 51.5 ± 13.5 years, NR: 45.0 ± 13.1 years), had higher BMI (PR: 28.4 ± 4.6 kg/m^2^, NR: 26.2 ± 4.4 kg/m^2^), were less likely to smoke (smokers PR: 6.5%, NR: 33.2%), and displayed slightly higher systolic and diastolic blood pressure (SBP and DBP respectively) (SBP PR: 127 ± 19.0 mmHg, NR: 121 ± 19.7 mmHg; DBP PR: 80 ± 10.8 mmHg, NR: 78 ± 11.8 mmHg) compared to individuals from the NR group (**Table EV1**). Dietary groups had similar distributions of female and male participants (female participants PR: 54%, NR: 55%), who were from families living above the line of poverty, were mostly married (PR: 73.5%, NR: 65.9%), had tertiary education (PR: 67%, NR: 75.4%), and originated mostly from Northern Greece (PR: 68.5%, NR: 65.4%) (**Table EV1**). Levels of physical activity were similar between groups and time points and the vast majority of participants were apparently healthy (according to self-reports), although a minority reported underlying chronic conditions, including diabetes, arterial hypertension and hypothyroidism, for which treatment was being received (**Appendix Table S1**). Very few participants were taking dietary supplements (**Appendix Text S3**).

We measured 16 blood biomarkers and 23 complete blood count (CBC) traits for the total of participants (N=411). Mean values of clinical chemistry measurements for each time point are summarised in **Table EV2**. For a subset of participants (PR N=28, NR N=19, **Appendix Table S2**) we isolated peripheral blood mononuclear cells (PBMCs) and performed immune cell profiling as well as stimulation experiments to investigate cell responses to immune challenges.

### Changes in biomarker levels, CBC traits and blood pressure

We found that short-term DR of animal products resulted in a notable shift in blood biomarkers, with almost all changes being in a direction that points to beneficial effects on health (**Table 1**, **Fig. 2**). Substantial reductions were found for levels of total cholesterol, low-density lipoprotein (LDL), urea, creatinine, alanine aminotransferase (ALT), and γ-GT. We also uncovered a striking 73% decrease in normal-range levels of CRP, and a very small, but significant reduction in BMI, after correction for multiple testing. Similarly to other lipoproteins quantified, we also found a small decrease for levels of high-density lipoprotein (HDL). Additionally, alkaline phosphatase (ALP) levels, that reflect bone turnover and liver health, were increased upon DR. In the control group, we only found small reductions in levels of creatinine and γ-GT from T1 to T2, possibly as a consequence of awareness bias following receipt of exam results at T1.

**Figure. 2.**
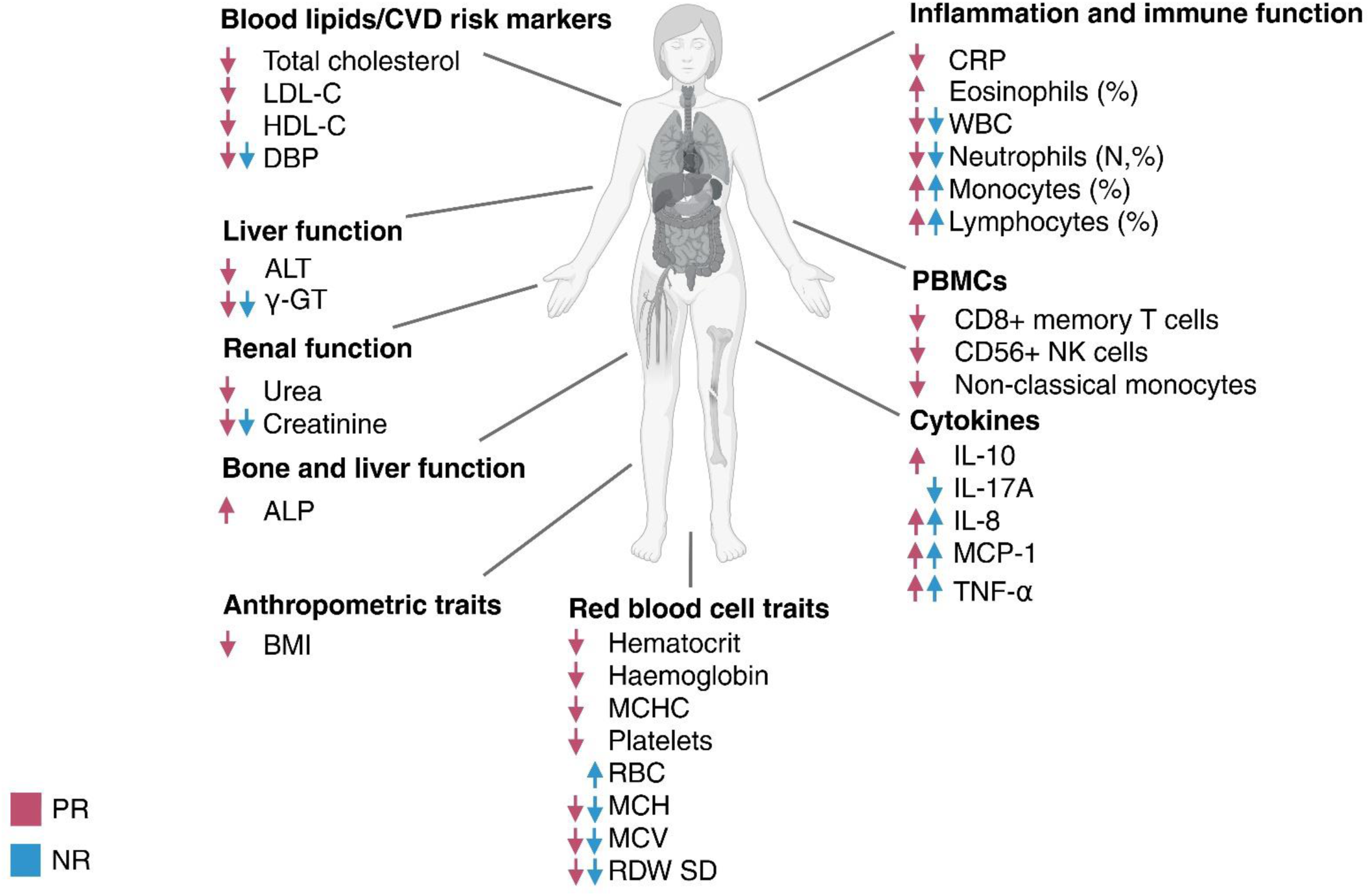
Effects of short-term dietary restriction of animal products. Changes in measured traits associated with short-term animal product restriction (PR individuals, red) were mostly beneficial for health. Fewer changes, likely capturing seasonal effects, were found in the continuously omnivorous group (NR individuals, blue) from T1 to T2.

**Table 1.**
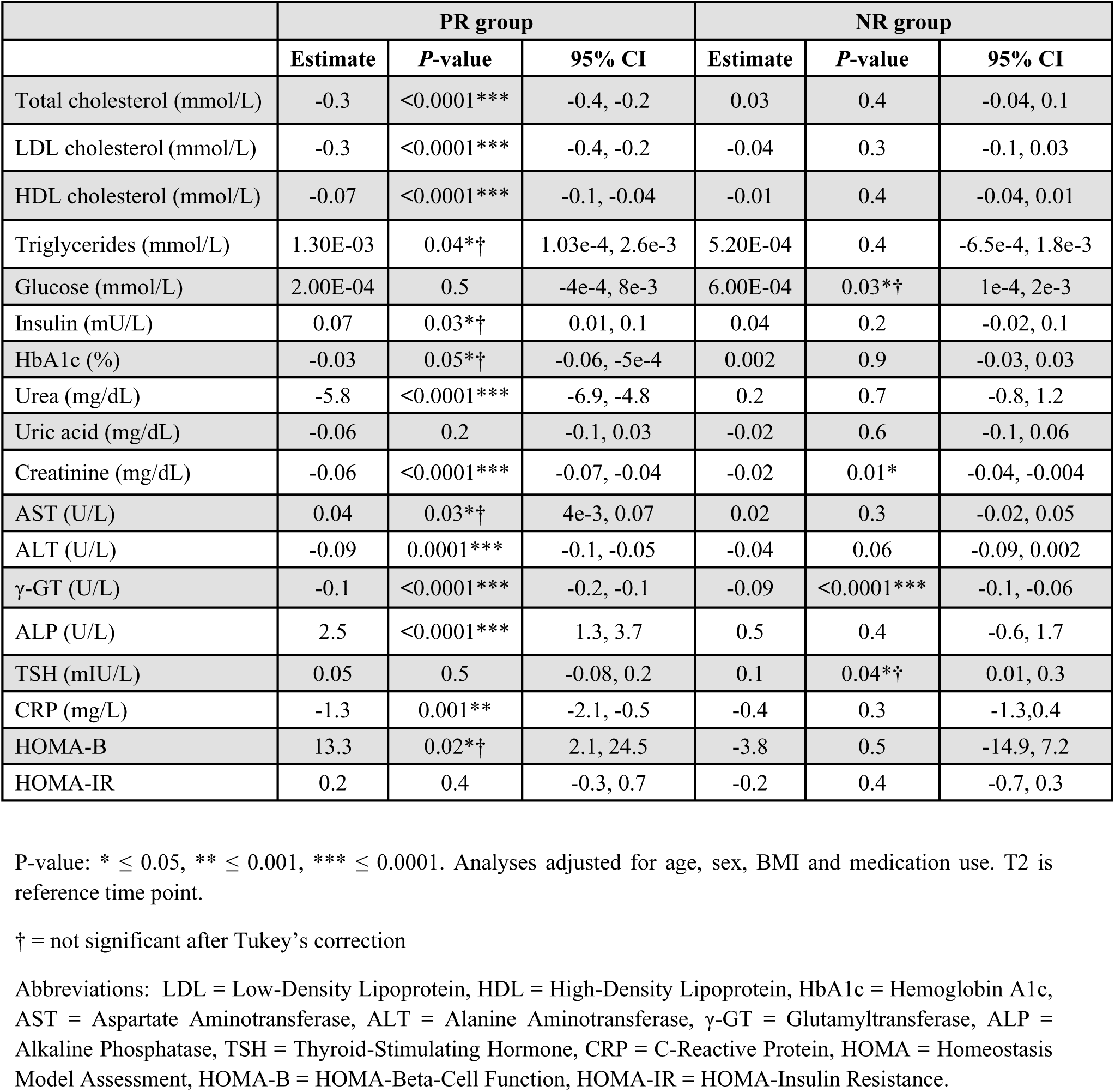
Changes in levels of blood biomarkers and of HOMA indices from T1 to T2.

CBC traits were also affected by DR with small reductions found for hemoglobin levels and mean corpuscular hemoglobin concentration (MCHC), haematocrit, and platelets **(Table 2**, **Fig. 2)**. An increase in eosinophil percentages was observed exclusively for the PR group upon DR. Both groups displayed a reduction in total white blood cell (WBC) count at T2. Although lymphocyte numbers did not change, lymphocyte percentages increased in both groups. Furthermore, at T2 we found significant reductions of neutrophil numbers and percentages, consistent with known seasonal effects for these cells (Wyse et al, 2021). Monocyte numbers and percentages increased in both groups at T2 (although the differences in monocyte counts were not significant following Tukey’s correction for the PR group). Both groups also displayed changes in red blood cell traits (mean corpuscular hemoglobin (MCH), mean corpuscular volume (MCV), and red cell distribution width standard deviation (RDW SD)), likely due to seasonal effects (Dopico et al, 2015) **(Table EV2)**. In the NR group only, we found a small increase in red blood cell counts (RBC) **(Table EV2)**. Furthermore, both groups displayed a small decrease in DBP **(Table EV3)**.

**Table 2.** Changes in complete blood count traits from T1 to T2.

|  | PR group |  |  | NR group |  |  |
| --- | --- | --- | --- | --- | --- | --- |
|  | Estimate | P-value | 95% CI | Estimate | P-value | 95% CI |
| WBC (K/μL) | -0.4 | <0.0001*** | -0.6, -0.3 | -0.3 | 0.0001*** | -0.5, -0.2 |
| Lymphocytes (x10<sup>3</sup>/μL) | -0.04 | 0.1 | -0.09, 0.01 | -0.008 | 0.8 | -0.06, 0.04 |
| Lymphocyte % (%) | 1.6 | 0.0004** | 0.7, 2.5 | 1.6 | 0.0003** | 0.8, 2.5 |
| Eosinophils (x10<sup>3</sup>/μL) | 0.0006 | 0.4 | -0.008, 0.02 | -0.01 | 0.07 | -0.03, 0.001 |
| Eosinophil % (%) | 0.2 | 0.02* | 0.05, 0.4 | -0.04 | 0.7 | -0.2, 0.2 |
| Monocytes (x10<sup>3</sup>/μL) | 0.02 | 0.02*† | 0.003, 0.03 | 0.02 | 0.005* | 0.007, 0.04 |
| Monocyte % (%) | 0.7 | <0.0001*** | 0.5, 0.9 | 0.6 | <0.0001*** | 0.4, 0.8 |
| Basophils (x10<sup>3</sup>/μL) | 0.0007 | 0.7 | -0.003, 0.004 | -0.0004 | 0.8 | -0.004, 0.003 |
| Basophil % (%) | 0.01 | 0.5 | -0.02, 0.05 | -0.01 | 0.4 | -0.05, 0.02 |
| Neutrophils (x10<sup>3</sup>/μL) | -0.4 | <0.0001*** | -0.5, -0.2 | -0.3 | <0.0001*** | -0.4, -0.2 |
| Neutrophil % (%) | -2.5 | <0.0001*** | -3.5, -1.6 | -2.1 | <0.0001*** | -3.05, -1.2 |
| RBC (M/mL) | 0.004 | 0.8 | -0.02, 0.03 | 0.07 | <0.0001*** | 0.04, 0.09 |
| Hemoglobin (g/dL) | -0.2 | <0.0001*** | -0.3, -0.1 | -0.02 | 0.6 | -0.1, 0.06 |
| Hematocrit (%) | -0.4 | 0.0003** | -0.7, -0.2 | 0.01 | 0.9 | -0.2, 0.2 |
| MCH (pg) | -0.5 | <0.0001*** | -0.6, -0.4 | -0.4 | <0.0001*** | -0.5, -0.4 |
| MCHC (g/dL) | -0.2 | 0.0001*** | -0.2, -0.08 | -0.07 | 0.08 | -0.2, 0.008 |
| MCV (fl) | -1 | <0.0001*** | -1.2, -0.8 | -1.2 | <0.0001*** | -1.3, -1.0 |
| RDW CV (%) | -0.08 | 0.05*† | -0.2, -4e-5 | -0.1 | 0.004*† | -0.2, -0.04 |
| RDW SD (fl) | -0.6 | <0.0001*** | -0.8, -0.3 | -0.8 | <0.0001*** | -1.0, -0.5 |
| Platelets (K/μL) | -9.8 | <0.0001*** | -13.4, -6.3 | -3.06 | 0.09 | -6.6, 0.4 |
| PDW (%) | 0.004 | 1 | -0.3, 0.3 | 0.02 | 0.9 | -0.2, 0.3 |
| MPV (μm) | -0.07 | 0.4 | -0.2, 0.09 | -0.04 | 0.7 | -0.2, 0.1 |
| P-LCR (%) | -0.9 | 0.006*† | -1.6, -0.3 | -1.2 | 0.0003**† | -1.9, -0.6 |

P-value: \* $\leq 0.05$ , \*\* $\leq 0.001$ , \*\*\* $\leq 0.0001$ . Analyses adjusted for age, sex, BMI and medication use. PR is reference group.
† = not significant after Tukey's correction
Abbreviations: WBC = White Blood Cells, RBC = Red Blood Cells, MCH = Mean Corpuscular Hemoglobin, MCHC = Mean Corpuscular Hemoglobin Concentration, RDW CV = Red Cell Distribution Width Coefficient Of Variation, Red Cell Distribution Width Standard Deviation = RDW SD, PDW = Platelet Distribution Width, MPV = Mean Platelet Volume, P-LCR = Platelet-Large Cell Ratio.

We also explored how the two dietary groups differed at each time point and found a striking resemblance at T1 when both groups were omnivorous (**Tables EV4, EV5, EV6**), with the only difference detected being lower eosinophil counts and percentages in PR individuals. This was not observed at T2 however, where we found substantial differences between groups, including lower levels of total and LDL cholesterol, urea, creatinine, and of WBC and neutrophil counts in PR individuals, but higher levels of ALP.

### Changes in cardiovascular disease risk profiles

DR was associated with a 0.3 mmol/L decrease in non-high-density-lipoprotein (non-HDL) cholesterol levels (p < 0.0001) in the PR group, while no change was observed in the NR group (β = 0.05, *P* =0.2). Non-HDL cholesterol is a comprehensive marker for assessing cardiovascular risk and reflects the total concentration of proatherogenic lipoproteins which contribute to atherosclerotic CVD (Hansen et al, 2024). This reduction in non-HDL was accompanied by a small reduction in LIFE-CVD2 risk score (calculated for participants > 35 years old, PR N=152, NR N=135) (Hageman et al, 2024) in the PR group (PR mean age = 54.0 years old, change in CVD risk score from 15.7% to 15.3%, p = 0.04), while no change was found for NR individuals (NR mean age = 51.0 years old, change in CVD risk score from 17.3% to 17.1%, p = 0.5).

### Ex vivo quantification of immune cells

We explored effects of animal product restriction on PBMC cellular composition using flow cytometry in a subset of individuals (PR N=28, NR N=19). Participant characteristics for the PBMC subgroup are shown in **Appendix Table S2**. Although small differences in viability were observed across dietary groups and time points, the overall viability of cells was over 75% in all cases. We found high interindividual variation in frequencies of major immune cell types for both dietary groups (**Fig. 3**), similar to the findings of a recent study on six individuals (Link et al, 2024). Although overall cell type proportions remained relatively stable between time points for each individual, we found that DR was associated with a reduced frequency of non-classical monocytes (*P* = 0.03), CD56^+^ NK cells (*P*=0.03), CD8^+^ memory T cells (*P*=0.0033) and a trend toward decreased frequency of CD8^+^ T cells (*P*=0.063). No changes were observed in the control group **(Fig. 4)**.

**Figure. 3.**
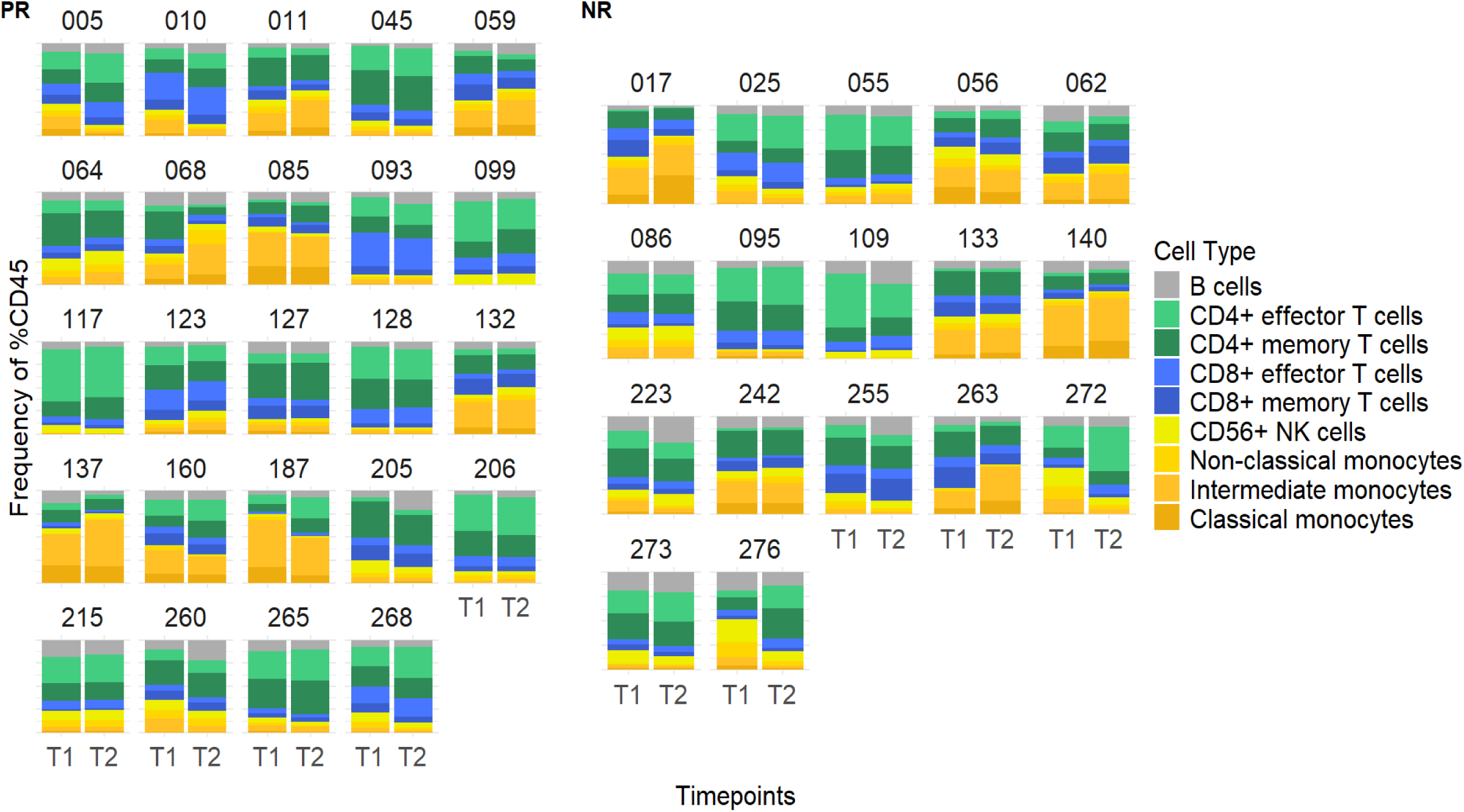
Distribution of immune cell types for PR and NR individuals at T1 and T2. Frequency of immune cell types (as frequency of CD45^+^ live cells) measured by flow cytometry. Numbers on top of mosaic plots represent participant IDs. Four PR and 2 NR individuals are not shown as CD4^+^ and CD8^+^ staining did not work.

**Figure. 4.**
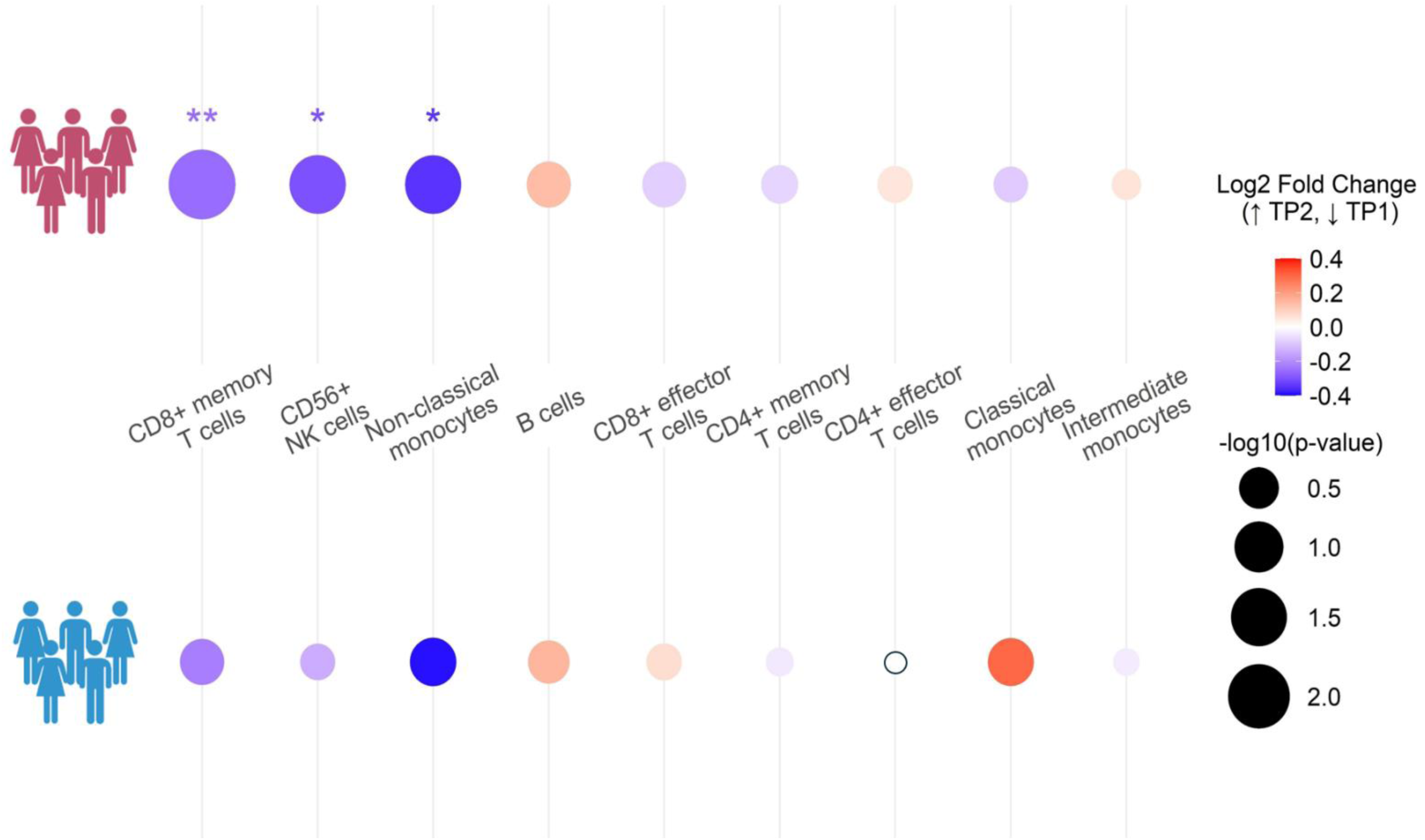
Fold change of cell populations whose frequency changed significantly between T1 and T2. Significant changes are indicated by *P* * < 0.05 and ** < 0.01. Significance was calculated by two-sided paired *t-*tests. Dots are scaled by -log_10_*P*. (Blue: reduced cell frequency at T2; red: increased cell frequency at T2). Exact *P*-values are mentioned in the text.

### Effects of animal product restriction on innate immune responses

We investigated the impact of animal product restriction on innate immune activation to the most common pathogen infections, by measuring cytokine levels produced by PBMCs after stimulation with lipopolysaccharide (LPS) or resiquimod (R848) for six hours. For all conditions investigated (unstimulated, LPS stimulation, R848 stimulation), no differences were found in PBMC viability, as measured through ATP quantification, the only exception being a slight decrease in ATP levels following LPS stimulation in the PR group (*P*=0.049). We calculated the magnitude of response to stimulation (log_2_ ratio of cytokine levels at stimulated vs unstimulated conditions) and found a trend for a mild response to LPS stimulation for IL-12p70 (*P*=0.053) **(Fig. 5A, Appendix Table S3)** following DR. Additionally, both dietary groups showed greater response to stimulation with LPS and R848 of pro-inflammatory cytokines IL-8, MCP-1 and TNF-α at T2 compared to T1 (*P*-value range 5×10^−5^-0.03) **(Figs. 5A and 5B, Appendix Table S3)** pointing to likely seasonal differences in the magnitude of response of innate immune cells to infection.

**Figure. 5.**
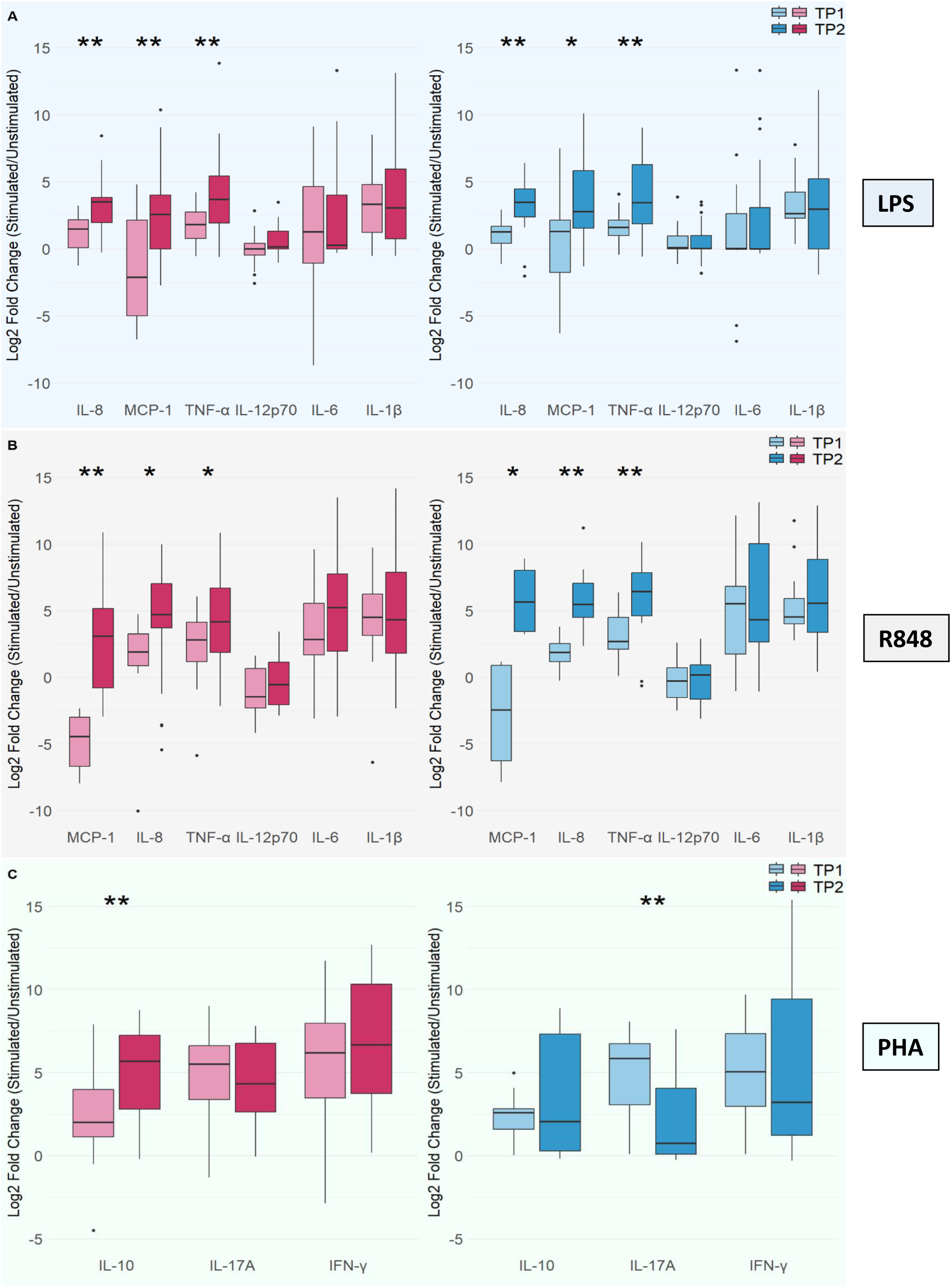
Magnitude of change in cytokine production between T1 and T2 following immune cell stimulation. **A.** Magnitude of change in cytokine production between T1 and T2 following 6-hour stimulation with bacterial ligand LPS. **B.** Magnitude of change in cytokine production between T1 and T2 following 6-hour stimulation with viral ligand R848. **C.** Magnitude of change in cytokine production between T1 and T2 following 48-hour stimulation with PHA. Significant changes are indicated by *P* * < 0.05 and ** < 0.01. Significance was calculated by two-sided paired *t-*tests. Exact *P*-values are provided in Appendix Table S3.

### Effects of animal product restriction on adaptive immune responses

We next examined the impact of animal product restriction on adaptive immunity by measuring cytokine levels produced by PBMCs following stimulation with T cell-selective mitogen phytohemagglutinin (PHA) for 48 hours. In individuals undergoing DR, we found a greater response for the anti-inflammatory cytokine IL-10 **(Fig. 5C, Appendix Table S3).** Notably, in NR individuals we found lower production of IL-17A at T2 compared to T1, in contrast to PR individuals whose ability to mount an IL-17A response at T2 was not compromised.

## Discussion

In the present study we explored effects of periodic short-term animal product restriction in a unique population sample from Greece who practice this dietary pattern for religious reasons for 180-200 days annually, and compared them to a continuously omnivorous control group. We found notable changes in blood biomarkers resulting from short-term DR, that are associated with positive effects on health, including a substantial decrease in levels of total and LDL cholesterol, improvements in markers of renal and liver function, and a striking 73% decrease in normal-range concentration of CRP. Furthermore, CVD risk profiles, as evaluated through non-HDL cholesterol levels and LIFE-CVD2 risk scores, were improved upon DR. These substantial changes were found to be rapid, occurring after three to four weeks of DR. However, the almost similar profiles of the two dietary groups when both were omnivorous suggests effects that are mostly transient. Both PR and NR groups showed reduced γ-GT levels at T2, in part likely reflecting lower alcohol consumption. PR individuals typically consume less alcohol during periods of DR (**Appendix Text S2**), while NR individuals possibly consumed less alcohol prior to T2 due to awareness bias at the second profiling time point.

Despite the short duration of DR, the magnitude of reductions in total and LDL cholesterol blood concentrations (0.3 mmol/L) was comparable to, or greater than, changes resulting from similar dietary interventions lasting three times longer (Rees et al, 2021; Termannsen et al, 2022). To put our findings in context, statin use reduces LDL cholesterol levels by an average of 1.8 mmol/L, reducing the risk of ischemic heart disease and of stroke by 60% and 17% respectively (Law et al, 2003). While the effects in our study are smaller, the magnitude of change is clinically relevant, suggesting that short-term DR of animal products can be applied as a complementary approach with rapid effects, to lipid-lowering therapies. Larger studies however, that investigate multiple outcomes over time are necessary to further support this idea.

In addition to reductions in urea and creatinine levels following DR, that likely reflect reduced protein intake (Georgakouli et al, 2022; Sarri et al, 2004), we found reductions in levels of ALT, suggesting reduced deposition of fat in the liver and to potential protection against hepatic steatosis (Despres et al, 1991; Sanyal et al, 2010; Straznicky et al, 2012). Furthermore, this short-term course of DR led to a 73% reduction in normal-range levels of CRP, suggesting a tempering effect of this dietary intervention on inflammation. Profiling of 1,464 plasma proteins in the same individuals has shown that short-term DR of animal products drives reductions in the levels of key proteins that are involved in inflammatory processes including HAVCR1, HPGDS, and PLA2G1B (Rouskas et al, 2025). HAVCR1 is a regulator of immune cell activity and of renal regeneration, and has been shown to increase the immune response by promoting activation and proliferation of T cells and cytokine secretion. PLA2G1B is a secreted phospholipase A2, while HPGDS catalyses the conversion of PGH2 to PGD2, a prostaglandin that acts as an early-phase mediator of inflammation. PLA2G1B and HPGDS are both involved in the production of inflammatory prostaglandins through the arachidonic acid pathway, and their reduced levels likely results in a decreased prostaglandin production and a tempering effect on inflammation. Given their role in promoting the release of pro-inflammatory cytokines and subsequent CRP production, we suggest that the decrease in CRP levels detected in the present study may be driven in part through effects of DR on the above proteins.

In addition to the reduction in CRP levels, we also found a reduction in platelet numbers upon DR. Both CRP and platelets have key roles in the formation of atherosclerotic lesions. CRP, along with other inflammatory markers, including IL-6 and IL-1β, is emerging as a potent target for atheroprotection (Ridker, 2021). Platelets constitute initial components in lesion development by binding to leukocytes and endothelial cells, and by initiating CD16^+^ monocyte differentiation into macrophages (Idzkowska et al, 2015; Lindemann et al, 2007). Our findings suggest that the potential atheroprotective effects of animal product DR may be mediated in part through reductions in the levels of CRP and platelets.

The present study has also revealed effects of DR on circulating immune cell subsets. Not surprisingly, we found high interindividual variation in immune cell frequency between participants, while cell frequencies between time points for each participant remained relatively unchanged. Importantly, PR individuals showed a significant reduction in the frequency of non-classical monocytes, whereas no changes were found in the NR group. Consistent with this finding, proteomic profiling of individuals in this population sample revealed increased abundance of CX3CL1 in PR individuals upon DR (Rouskas et al, 2025). CX3CL1 is a chemokine that promotes migration of immune cells, including monocytes, from the blood into tissues (Lee et al, 2018). Monocytes are crucial for the induction and maintenance of inflammation, with CD16^+^ non-classical subsets being the most pro-inflammatory and senescent subset compared to classical and intermediate monocytes (Ong et al, 2018; Stansfield & Ingram, 2015). Senescent cells accumulate with age (Tominaga, 2015) and contribute to chronic low-grade inflammation, driving inflammaging, which is pivotal for the development and progression of age-related conditions, including atherosclerosis, osteoarthritis, cardiovascular and neurodegenerative diseases, type 2 diabetes mellitus, and cancer (Fulop et al, 2016; Tizazu, 2024). DR-associated decreased frequency of circulating non-classical monocytes may thus contribute to an overall improved inflammatory profile, as shown by the decreased normal-range CRP levels and the stronger response of the anti-inflammatory cytokine IL-10. Under conditions of caloric restriction, Jordan et al found that the number of blood monocytes decreased, and their metabolic activity changed (Jordan et al, 2019). Although the dietary pattern practiced by PR individuals does not involve a decrease in total energy intake (Georgakouli et al, 2022; Sarri et al, 2004), it may engage overlapping pathways given that caloric restriction involves the restriction of all macronutrients. Taken together, these data indicate that animal product restriction may suppress systemic inflammation, potentially contributing to better outcomes in chronic inflammatory diseases (Choi et al, 2017).

In addition to non-classical monocytes, blood CD56^+^ NK cells and CD8^+^ memory T cells, which both have cytotoxic function, were reduced during animal product restriction. It is conceivable that this reduction in circulating cytotoxic cells may negatively impact cell-mediated immunity against microbial infection. However, recent evidence showed that NK and memory T cells vanished from the blood and found residence in the bone marrow (BM) during caloric restriction in mice and humans (Collins et al, 2019; Delconte et al, 2024). Similarly to monocytes, CX3CL1 has been found to mediate chemotaxis of T cells into the BM in humans (Ren et al, 2014). In addition, nutrient restriction enhanced the metabolic fitness of cytotoxic cells, protecting them from exhaustion. BM provides a safer microenvironment that may better protect long-lived immune cells from nutrient deprivation in the blood and preserve their function. Although we cannot currently exclude that blood NK and memory CD8^+^ T cells may undergo apoptosis during animal product restriction, it is possible that animal product DR may drive relocation of immune cells away from the circulation, thus prioritizing cytotoxic memory and innate-like responses under nutrient stress, while dampening down energy-costly inflammation. These results suggest the intriguing possibility that timed DR may be used as a means to alter immune responses to boost anti-microbial or anti-tumoral immunity or vaccine responses, or to mitigate the risk of autoimmune or chronic inflammatory diseases.

Importantly, our study captured seasonal effects on the immune system, with both dietary groups exhibiting changes in WBC counts, lymphocyte percentages, monocyte percentages, neutrophil counts, and neutrophil percentages, in concordance with findings from the UK Biobank (Wyse et al, 2021). Likely seasonal changes were also found for both dietary groups for innate immunity pro-inflammatory cytokines IL-8, MCP-1 and TNF-α.

Overall, the majority of changes detected upon DR were in a direction associated with beneficial effects on health. However, we found that ALP levels increased, suggesting possible negative effects on bone or liver health (Kuo & Chen, 2017). Individuals who follow a vegan diet have been shown to have a higher risk for bone fractures (Tong et al, 2020) and we suggest that higher ALP levels may reflect negative effects of periodic animal product DR on bone health. Indeed, our work on plasma proteomics has shown DR-associated changes in levels of OXT, SPP1, FGF23, PTH1R and PTHrP, in a direction pointing to detrimental effects on bone homeostasis (Rouskas et al, 2025). Further research using more representative tests such as quantification of bone mineral density (BMD), bone-specific ALP, C-terminal telopeptide (CTx) and parathyroid hormone is needed, to better understand effects of this dietary pattern on bone health (Greenblatt et al, 2017).

Our study points to rapid, largely beneficial effects of short-term animal product restriction on health. Our work however, is limited by the fact that most blood biomarkers measured are known to capture relatively acute dietary effects, rather than detecting potential long-term effects on human health. Additionally, we have measured inflammation through quantification of CRP levels instead of high-sensitivity CRP (hs-CRP), which is more commonly used to detect the risk for inflammatory conditions (Lee & Lee, 2023). Although we have adjusted for confounding factors to the best of our ability, some residual effects may persist in reported results. Finally, cytokine responses were measured in total PBMCs meaning that we cannot pinpoint responses to specific cell types. There are also caveats to utilizing cryopreserved PBMCs which have resulted in loss of the more fragile cell subsets and overall loss of immune function. Despite these limitations, our study’s findings offer valuable insights for clinical practice and future intervention strategies using less restrictive plant-based diets which have likely beneficial effects on cardiometabolic and immune system health.

## Methods

### Recruitment of participants, collection of data and of biological material and measurement of traits

We screened over 1,000 candidate participants from the greater region of Thessaloniki, Greece for the FastBio (religious <u>Fast</u>ing <u>Bio</u>logy) study (Rouskas et al, 2025). Following two interviews, 411 apparently healthy, unrelated individuals, who fulfilled the selection criteria (**Appendix Text S1**) were included in the study. Participants belonged to one of two groups, specified by their diet: 200 individuals who followed a temporally structured dietary pattern of animal product restriction (periodically restricted, PR group) and 211 continuously omnivorous individuals who follow the diet of the general population and were not under any kind of special diet (non-restricted group, NR group). PR individuals constitute a unique study group given their consistent adherence to the dietary pattern specified by the Greek Orthodox Church. This pattern involves alternating between omnivory and restriction of animal products, including meat, fish, dairy products and eggs (but not molluscs and shellfish), for 180-200 days annually (**Appendix Text S2**). DR is practiced during four extended periods throughout the year, and on Wednesdays and Fridays of each week (Rouskas et al, 2025). Only PR individuals who had practiced this dietary pattern for at least ten years were included in the study. During periods of restriction, PR individuals also typically consume less alcohol. Although practicing individuals undergo restriction of animal protein intake, total energy intake is not reduced (Georgakouli et al, 2022; Sarri et al, 2004). NR participants were continuously omnivorous individuals from the general population, who were not under any kind of special diet.

FastBio participants were invited to two scheduled appointments at the Interbalkan Hospital of Thessaloniki. The first appointment took place in October-November 2017 (time point 1, T1) and covered a period where all PR individuals had been on an omnivorous diet (excluding Wednesdays and Fridays) for a period of eight or nine weeks. The second appointment (time point 2, T2) took place in March 2018, and covered a period during which PR individuals had abstained from meat, fish, dairy products and eggs for three to four weeks during the abstinence period prior to Easter (Lent). At T2, a recall rate of 95% was achieved, with 192 (out of 200) PR participants and 198 (out of 211) NR participants attending both appointments. All appointments were scheduled between 7:30-9:30 am to minimize circadian effects and were completed during a two-week window to minimize effects of seasonality. The ethics committee of BSRC Alexander Fleming approved the study protocols, the study was performed in line with the principles of the Declaration of Helsinki, and all participants provided written informed consent.

For each participant, following overnight fasting, a blood sample was collected by a certified nurse and was taken to clinical chemistry measurements (biomarker quantification and CBC testing) at the InterBalkan Hospital facilities (cobas c 311 analyzer, Roche Diagnostics). For a subset of 47 participants (PR N=28, NR N=19) an additional 20 mL of blood was drawn for isolation of PBMCs.

We profiled 16 blood (serum) biomarker traits and 23 CBC traits (**Table EV2**). Biomarkers were grouped to allow for broad interpretation (Tong et al, 2021): blood lipids (total cholesterol, LDL cholesterol, HDL cholesterol, triglycerides), glucose metabolism (glucose, insulin, hemoglobin A1c (HbA1c)), renal function (urea, uric acid, creatinine), liver function (aspartate aminotransferase (AST), ALT, γ-GT), bone and liver function (ALP), thyroid function (thyroid-stimulating hormone (TSH)), and inflammation (CRP). Using glucose and insulin measurements, we calculated homeostatic model assessment (HOMA) indices to assess beta-cell function (HOMA-B) and insulin resistance (HOMA-IR) **(Appendix Text S4)**. Height was measured to the nearest 0.5 cm using a stadiometer. Body weight was measured with the use of a calibrated digital scale with an accuracy of ±100g. BMI was calculated as body weight divided by the square of height (kg/m^2^). SBP and DBP were measured using a traditional sphygmomanometer.

### Peripheral Blood Mononuclear Cell Isolation

PBMCs were isolated from 20 mL of peripheral venous blood using density gradient centrifugation (standard procedure). In brief, blood was centrifuged at 1,500 rpm for 5 minutes. After plasma was removed, the remaining fraction was first diluted with an equal volume of DMEM (Gibco, Grand Islands, NY, USA) and then overlaid on Histopaque (specific gravity 1.077 g/mL; Sigma-Aldrich, St Louis, MO, USA) and finally centrifuged at 4,000 rpm for 30 minutes. PBMCs were isolated from the mononuclear cell layer, aliquoted into cryovials and stored in liquid nitrogen for subsequent experiments.

### Flow cytometry

Thawed PBMCs were transferred into a 15 mL conical tube containing 10 mL media (10% FBS, 1% Pen/Strep, RPMI 1640). The suspension was centrifuged at 330xg for 10 minutes at room temperature, decanted and resuspended in 1 mL media for counting of viable cells with an automated cell counter and trypan blue staining (Corning 100mL Trypan Blue Solution, 0.4% (w/v) in PBS, #25-900-CI; CellDrop, DeNovix). To determine the impact of animal product restriction on immune cell populations, we performed multiparametric flow cytometry to identify immune cell subsets. Five hundred thousand live cells were seeded into 275 µL media. One mL 1x PBS was added to acclimated cells and centrifuged at 330xg for 10 minutes at room temperature. Cells were stained with Zombie Green^TM^ (Fixable Viability Kit, BioLegend, #423111) at a 1:100 dilution in 1x PBS and incubated for 30 minutes. Fc receptor blocking (anti-CD16/CD32) was performed in fluorescence-activated cell sorting (FACS) buffer for 10 minutes at room temperature. PBMCs were then stained with surface antibodies (BioLegend and ThermoFisher) grouped into myeloid and lymphoid panels **(Appendix Table S4).** To minimize batch effects, we ran samples from both dietary groups and both time points on the same run, with three participants per run for a total of 16 runs.

Nine non-overlapping cell types were presented for analysis and interpretation: classical monocytes (CD45^+^/CD11b^+^/CD14^+^CD16^−^), non-classical monocytes (CD45^+^/CD11b^+^/CD14^−^ CD16^+^), intermediate monocytes (CD45^+^/CD11b^+^/CD14^+^CD16^+^), CD4^+^ effector T cells (CD45^+^/CD66b^−^CD14^−^/CD3^+^CD19^−^/CD4^+^CD8^−^/CD45RO^−^), CD4^+^ memory T cells (CD45^+^/CD66b^−^CD14^−^/CD3^+^CD19^−^/CD4^+^CD8^−^/CD45RO^+^), CD8^+^ effector T cells (CD45^+^/CD66b^−^CD14^−^/CD3^+^CD19^−^/CD4^−^CD8^+^/CD45RO^−^), CD8^+^ memory T cells (CD45^+^/CD66b^−^CD14^−^/CD3^+^CD19^−^/CD4^−^CD8^+^/CD45RO^+^), B cells (CD45^+^/CD66b^−^CD14^−^/CD3^−^CD19^+^) and CD56^+^ NK cells (CD45^+^/CD66b^−^CD14^−^/CD3^−^CD19^−^/CD56^+^). Stained cells were acquired using the Novocyte flow cytometer (Agilent) and analysed using Flowjo v10.7.2 (BD) **(Appendix Figs. S1 and S2, Appendix Table S5).** CD4^+^ and CD8^+^ staining did not work in 4 PR and 2 NR individuals, therefore these samples were excluded from the analysis for these specific cell types, and they are not shown in **Fig. 3**.

### PBMC stimulation experiments

To explore whether animal product restriction has an impact on cytokine production in PBMCs, cytokine production was measured *in vitro* upon stimulation with LPS or R848 for 6 hours, or with PHA for 48 hours. For the 6-hour stimulation experiments, 100,000 live cells were seeded into 96-well round-bottom wells with 200 µl media per well. Cells were left in media alone (control) or stimulated with Toll-like receptor (TLR)-ligands: LPS (100 ng/mL), or R848 (10 µg/ml), in technical triplicates. Subsequently, the supernatant was collected for cytokine quantification with 13-plex LEGENDplex^TM^ kit (BioLegend, Human Inflammation Panel 1 (13-plex) w/VbP, #740809) and 13-plex LEGENDplex^TM^ kit (BioLegend, Human Anti-Microbial Panel (13-plex) #714269), and cells assessed for viability using the Cell-Titer-Glo 2.0 Cell Viability Assay (Promega, #G9241). For the 48-hour stimulation experiments, 500,000 live cells were plated in 96-well round-bottom wells with 275 µL media per well. Upon plating, cells were stimulated with PHA (10 µg/ml) to assess T-cell activation and proliferation. The supernatants were collected for cytokine measurement using the 13-plex LEGENDplex^TM^ kit (BioLegend, Human Inflammation Panel 1 (13-plex) w/VbP, #740809). **(Appendix Table S6).**

### Statistical analysis of clinical chemistry measurements, anthropometric traits and blood pressure

We performed multivariable regression analysis using linear mixed effects (lme) models with random effects (“nlme” R version 4.1.1) to find changes in measured traits between time points, with T2 as the reference time point (**Appendix Text S5**). We also explored differences between dietary groups at each time point (PR as the reference dietary group). Tukey’s correction was performed for all associations and *P* < 0.05 were considered statistically significant. Eight non-normally distributed traits (triglycerides, glucose, insulin, creatinine, AST, ALT, γ-GT, and CRP) were log-transformed prior to regression analyses. For CRP, 49 individuals (25 PR and 24 NR) with values suggesting acute inflammation (CRP > 5 mg/L) were excluded from further analysis. The model adjustments for the clinical chemistry measurements and the anthropometric and blood pressure traits are described in **Appendix Text S5 and Appendix Table S7**. Individual heterogeneity was accounted for by including participant ID as a random intercept in the models.

### Calculation of non-HDL cholesterol and lifetime CVD risk profiles

We sought to assess CVD risk profiles based on non-HDL cholesterol levels, by deducting HDL cholesterol from total cholesterol for each participant (Hansen et al, 2024; Raja et al, 2023). Lifetime CVD risk was evaluated for participants > 35 years old, according to ESC guidelines, using the LIFE-CVD2 risk calculator (Hageman et al, 2024). We calculated the LIFE-CVD2 score for 152 PR individuals (81 females and 71 males) and 135 NR individuals (65 females, 70 males) at each time point. A Wilcoxon signed-rank test was used to assess change (%) in CVD risk score between time points and *P* < 0.05 were considered statistically significant.

### Statistical analysis of immune cell frequencies and PBMC cytokine measurements

We interrogated changes in cell frequencies between T1 and T2 for each dietary group through paired sample t-tests (R version 4.3.3). Results with *P* < 0.05 were considered statistically significant. For cytokines, we explored the magnitude of the difference between unstimulated and stimulated conditions (magnitude of response to stimulation) at each time point for each dietary group, by calculating the log_2_ ratio of cytokine levels at stimulated vs. unstimulated conditions (log_2_fold change for response to stimulation). We compared responses between time points for each dietary group through paired sample t-tests.

## Acknowledgements

The authors would like to thank the FastBio study participants and the Interbalkan Hospital staff. We would also like to thank Dr Dimitrios Rouskas, Dr Loukas Kipouros and Dr. Allen Seylani for their invaluable help. Finally, we would like to thank Professor Stylianos Antonarakis for his helpful comments on this work.

The FastBio study was supported through a European Research Council Grant to Antigone Dimas (FastBio – 716998). Research at the University of California, Riverside was supported by the School of Medicine.

## Data availability

The data underlying this article will be shared on reasonable request to the corresponding author. This study includes no data deposited in external repositories.

## Author contributions

EML: Conceptualization, Methodology, Formal analysis, Writing – original draft. AP: Methodology, Formal analysis, Writing – original draft. SA: Methodology, Formal analysis, Writing – original draft. AS: Formal analysis, Writing – Review & Editing. PB: Formal analysis, Writing – Review & Editing. SG: Data acquisition, Material Preparation, Writing – Review & Editing. AD: Data acquisition, Material Preparation, Writing – Review & Editing. MA: Data acquisition, Material Preparation, Writing – Review & Editing. KB: Data Acquisition, Material Preparation. IC: Formal Analysis, Interpretation. EK: Interpretation. PR: Interpretation, Writing – Review & Editing. IK: Conceptualization, Methodology, Interpretation, Writing – Review & Editing. ND: Conceptualization, Methodology, Interpretation, Writing – Review & Editing. NS: Conceptualization, Methodology, Interpretation, Writing – Review & Editing. MY: Interpretation, Writing – Review & Editing. MV: Interpretation, Writing – original draft. MGN: Conceptualization, Methodology, Writing – original draft. KR: Data acquisition, Material Preparation, Writing – Review & Editing. ASD: Supervision, Funding acquisition, Conceptualization, Methodology, Data acquisition, Material Preparation, Writing – original draft.

EML, AP, SA contributed equally to this work.

